# Designing a text messaging program to increase adherence to medication for the secondary prevention of cardiovascular disease

**DOI:** 10.1101/19002683

**Authors:** Ana Uribe-Rodríguez, Paula Pérez-Rivero, Caroline Free, Pablo Perel, Elizabeth Murray, Norma Serrano Díaz, Robert Horne, Louise Atkins, Juan Pablo Casas, Anderson Bermon Angarita

## Abstract

**Background:** Cardiovascular medication for secondary prevention has been shown to be effective. However, cardiovascular patients have poor medication adherence, the consequences of which include premature death, recurrence risk, hospitalization, and high financial cost for the healthcare system. Behavioral interventions based on text messaging technology are a promising strategy to improving adherence in medications. In low-middle income settings there is no high-quality evidence of a behavioral program delivered by SMS; hence we describe the development, message content, and the program design of the intervention for improving adherence to cardiovascular medication.

**Methods:** We used the model reported by Abroms and colleagues’ for developing and evaluating text messages-based interventions. This model describes a process in which the intervention created is based on theory and evidence, the target audience is involved to ensure the intervention is engaging and useful, and there is a focus on implementation from the outset.

**Results:** Our main result was the design of the program, which consisted of a twelve-month structured intervention based on Transtheoretical Model of Behavior Change. We wrote and validated clusters of texts messages targeting each stage of the model. Each message went through an examination process including the evaluation of former cardiovascular patients, experts and the team research personnel. Another important result was an understanding of patients’ perceptions of their experience of cardiovascular disease, barriers to accessing healthcare in Colombia and the use of mobile technology for health.

**Conclusions:** An SMS intervention has the potential to be an acceptable and effective way of improving adherence to medication in patients with cardiovascular disease. This paper describes the development and content of one such intervention.

## Background

People with a history of cardiovascular events have a five times higher risk of new events than people without no such history [1].Secondary prevention medications (anti-platelet therapy, ACE inhibitors, beta-blockers and lipid lowering therapy) are effective in reducing the risk of death and myocardial infarction in patients with coronary heart disease. Meta-analysis of randomized controlled trials show that in patients with existing coronary heart disease long term anti-platelet therapy reduces major vascular events (MI, stroke or vascular mortality) by about a quarter (OR 0.75 (95% CI 0.71-0.79); ACE inhibitors reduce cardiovascular mortality by just under a fifth (RR 0.83, 95% CI 0.72 to 0.96); beta blockers reduce mortality by almost a quarter (OR 0.77, 95% CI 0.69 to 0.85); and lipid lowering therapy reduces coronary mortality by about a fifth (RR 0.79, 95% CI 0.75 to 0.83) [2].

However, cardiovascular patients have poor medication adherence, the consequences of which include premature death, increased risk of further cardiovascular events, hospitalization, and high financial cost for the healthcare system [3] [4] [5] [6] [7]. Therefore, strategies to improve medication adherence to cardiovascular drugs must be a health priority [8] [6]. Since 2002, text messaging, or short message service (SMS), has been part of m-health strategies to improve health and change behavior [9]. Interventions using SMS have been shown to be effective for smoking cessation, adherence to antiretroviral therapy [10], diabetes self-management, weight loss, and physical activity [9]. As for cardiovascular treatment, several randomized clinical trials (RCT) using text messages to improve medical adherence in primary as well for secondary prevention have produced promising, although not yet conclusive, results [11] [12]. Systematic reviews conclude that existing RCTs have high risk of bias, so the evidence is uncertain, and most were performed in high-income countries, so the evidence may not generalize to low income countries [11].

The ongoing randomized clinical trial (RCT) “TXT2HEART COLOMBIA: Evaluation of the Efficacy and Safety of Text Messages to Improve Adherence to Cardiovascular Medications in Secondary Prevention” (https://clinicaltrials.gov), involves an SMS-based intervention aimed at improving adherence to cardiovascular medications amongst adults with history of arterial occlusive events living in a metropolitan area of Colombia. This paper presents the design and validation process of text messages developed for the RCT based on Transtheoretical Model of Health Behavior Change (TTM) [13]. This model, also known as the Stages of Change, integrates cognitive, behavioral, and motivational factors at different times in the behavior modification process. It has been recently applied to develop interventions that use SMS to change behavior related to nutrition [14], physical exercise [15] [16] and hypertension [17]. The publication of this phase of the study will contribute to filling gaps in the literature identified by Adler et al [11] by describing the development, message content, and the program design of the intervention. It also fulfils the requirement to publish detailed descriptions of interventions to avoid research waste [18] and allow a growing evidence base about which interventions work for what reason [19].

## Method

Our methodological approach is described in Fig. 1. We used the model reported by Abroms and colleagues’ [20] for developing and evaluating text messages-based interventions. This model describes a process in which the intervention created is based on theory and evidence, the target audience is involved to ensure the intervention is engaging and useful, and there is a focus on implementation from the outset. Each step will be described in this section.

**Figure 1.**
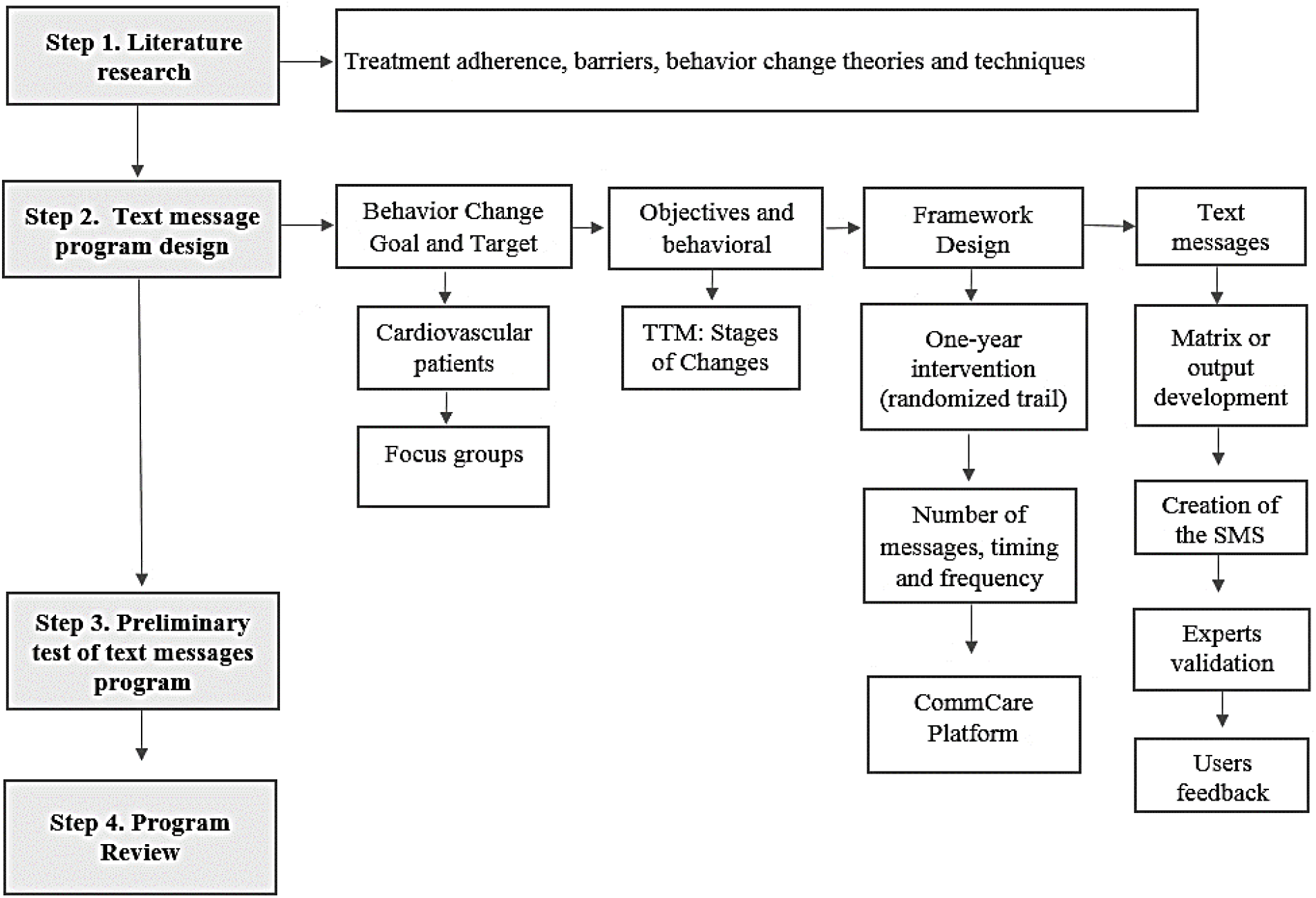
Process for text message intervention design.

Literature research: the first step consisted in conducting literature research related to the target audience and behaviour change theories. A successful health program needs evidence-based strategies [21] [22] and must focus on understanding the health behavior that characterizes the population [20]. Literature research aimed to: 1) understand the factors associated with low adherence to cardiovascular medication and 2) identify the leading models and appropriate techniques for modifying health behaviors related to medication adherence. We performed literature research from February 2015 to June 2016 in the following database: Scopus, Ovid, Medline, Cochrane Library, EBSCO, Pub-Med, Science Direct and Google Scholar. The keyword terms were: Medication adherence, Medication adherence and cardiovascular Disease, Social support and cardiovascular risk, Health models, and M-Health. We reviewed the following behavior change models: The Theory of Reasoned Action [23], Social Learning Theory [24], the Transtheoretical Model of Change [13], the Precede Model [25] and the Behavior Change Wheel [26].

Objectives and techniques: As a result of this literature review, three main objectives for the intervention were established based on the behavior change goal and the theoretical aspects of the TTM: (1) raising awareness about medication effectiveness, (2) promoting self-care, and (3) offering guidelines for behavioral strategies to increase adherence (see Table 2). When the TTM was first developed, Prochaska and DiClemente [13] proposed a relationship between the perception of illness, or more appropriately the rise of awareness of illness, and the intention to change behavior and the subsequent change.

Text message program design process: Our behavior change goal was to improve medication adherence in secondary prevention for patients with history of occlusive arterial disease. In the literature search we found several studies indicating the importance of evaluating barriers that affect adherence to medications in cardiovascular patients [5] [27] [28]. We carried out qualitative research to identify barriers to adherence in our setting and compared our findings to the existing literature.

We used focus group methodology to interview patients with a previous history of cardiovascular events. Participant inclusion criteria were: male and female adults (18 years and older), ownership of a mobile cell phone with an active line, ability to read text messages and to give informed consent, and at least a six-month history of any of the following cardiovascular diagnoses: coronary artery disease (including non-primary coronary revascularizations), ischemic stroke, and peripheral artery disease or atherosclerotic aortic disease. These selection criteria mirrored the eligibility criteria for the TXT2Heart trial. Recruitment focused on patients who had an occlusive arterial disease event and been hospitalized at the Fundación Cardiovascular FCV, which provided clinical evidence of the event as well easier access to the patients. The inclusion criteria were narrowed to events occurring between six months and five years prior to the study. Participants were randomly selected from the FCV databases using SQL (Structured Query Language), with specification of the date of the event (between July 2011 and July 2015), ICD 10 diagnosis, and medical procedure such as angioplasty, revascularization, and other interventions for the cardiovascular diseases mentioned. After the patients gave their consent, a trained physician reviewed the patients’ electronic clinical histories to confirm the diagnoses and dates of the occlusive event.

Text Message Library: The text message designers worked as an interdisciplinary team, which included an epidemiologist, a pharmacologist, a specialist nurse in health and quality audits, and three psychologists. Each team member developed messages according to the following recommendations: length up to 160 characters (including spaces), use of appropriate language, avoidance of abbreviations and misspellings, and informal punctuation [20]. Most importantly, for the messages content the team considerate the findings from the focus groups (categories and subcategories) and TTM (Stages of Change). Once the messages were written, the interdisciplinary team held several discussion meetings to evaluate and classify each text message, considering aspects such as message pertinence, intention, and clarity.

The messages were classified as viable, editable or as messages to be discarded. Those identified as editable were rewritten, and some were also eliminated. Then, the messages were organized using a matrix and classified according to the categories and subcategories related to the barriers to adherence that were identified in the literature and the focus groups. This helped to visualize how many text messages corresponded to each category and subcategory, and to develop new ones in order to have at least 12 messages in each category.

Expert validation: The text messages were submitted for evaluation by 13 experts. The panel consisted of several different types of health professionals, including psychologists (clinical, psychometric, and general), nutritionists, nurses (practitioner, administrative, and researcher) and physicians (general practitioner, cardiologist, interventional cardiologist, and with expertise in prevention and health promotion). The experts were selected through convenience sampling and the inclusion criteria were knowledge of the topic (cardiovascular disease for physicians and behavior change for psychologists), research experience, and test/trial validation expertise. These professionals were invited to participate through a letter that explained the purpose of the validation. After accepting, they received two documents: (1) instructions for validating the text messages, which included descriptions of the categories, stages of the TTM, and aspects to be evaluated (clarity, coherence, and relevance), and (2) an Excel template containing the text messages, the categories and subcategories, aspects to be evaluated, and an additional space to record observations. The psychologists participating as experts also evaluated the coherence between the SMS and the TTM of change.

All the judges evaluated the messages for clarity, coherence, and relevance on a scale of 1 to 4, with 4 as the best score. An average score of 3.7 to 4 for the three aspects and a Kendall’s W of .85 or higher were indicators of strong agreement among the judges.

Pretesting and revising the text messaging program: the third step included feedback from prospective users about messages presented to them face to face, international researchers’ opinions, and finally a test for the deliverability of messages to cell phones. To obtain feedback form users, the participants from previous focus groups were invited to attend a new focus group and to evaluate the text messages. A total of 9 cardiovascular patients attended the meeting. The focus group was facilitated by a psychologist and had four observers (one physician, one nurse, and two psychologists). Participants were asked to evaluate clarity, coherence (according to the categories), and the importance and relevance of the messages. The final messages were presented to international experts in the development of messaging interventions for comments. Lastly, a test was conducted to assess the delivery of the text messages program using *the CommCare* and *Telerivet* platforms. To this end, some of the text messages were sent to the research team’s cell phones.

## Results

This section describes the results from the different steps presented in the Methods section.

### Literature Search

The World Health Organization [29], listed five dimensions that affect treatment adherence: social and economic factors; health care and systems-related factors; condition related factors; therapy related factors; and patient related factors. The literature consistently shows that treatment adherence in cardiovascular patients is poor [3] [4] [5] [6] [7]. Authors describe the non-adherence of cardiovascular medication as a global threat [6] and as the major obstacle to the treatment success [3]. Poor adherence to medication appears to be only minimally related to the class of drugs taken or their side effects [4]; however, misinformation, lack of knowledge about, or awareness of, cardiovascular disease is an important barrier to treatment adherence [30] [7] [31].The asymptomatic nature of many cardiovascular diseases, as well, may be related to poor adherence to prescribed medication [23] [5]. As expected, difficulties with insurance coverage are associated with poorer cardiovascular health [32] while perceived social support can be related to better adherence [5].

In Latin America a study identified that lack of awareness of guidelines and knowledge about preventing cardiovascular disease, communication problems within health teams and lack of motivation were barriers to treatment adherence. Particularly in Colombia, studies with cardiovascular patients’ show that most participants are at risk of non-adherence [33].

### Focus groups

Forty cardiovascular outpatients were approached and invited to the focus groups. Participants ranged in age from 25 to 80 years and most were male. Data from participants were coded into the categories and subcategories described in Table 1.

**Table 1.**
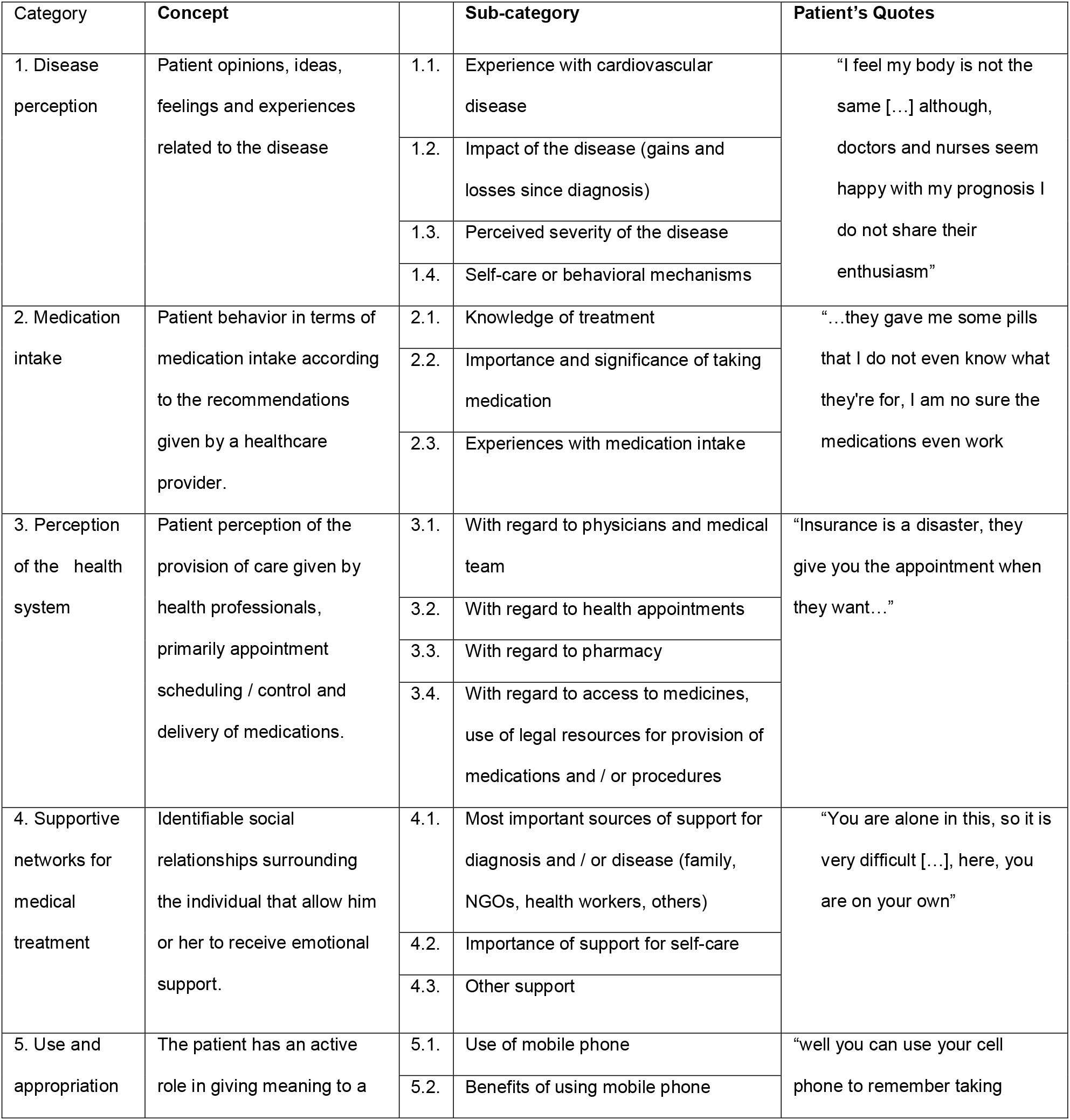
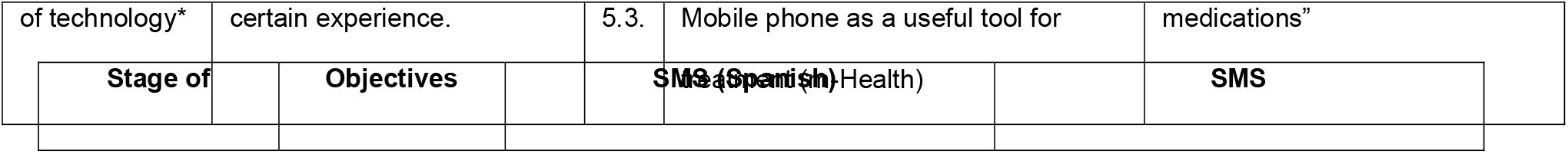
Categories and subcategories.

^1^ EPS- Common name in Colombia for the different companies that manage health insurance

### Objectives and Techniques

Three main objectives for the intervention were established based on the behavior change goal and the theoretical aspects of the TTM: (1) raising awareness about medication effectiveness, (2) promoting self-care, and (3) offering guidelines for behavioral strategies to increase adherence (see Table 2). When the TTM was first developed, Prochaska and DiClemente [13] proposed a relationship between the perception of illness, or more appropriately the rise of awareness of illness, and the intention to change behavior and the subsequent change. During this course of action, people experience different processes of change and progress through five stages: pre-contemplation, contemplation, preparation, action, and maintenance. These stages represent the temporal dimension of behavior change [34] and a shift from intention to action. The model includes at least ten processes of change (consciousness raising, dramatic relief, environmental re-evaluation, social liberation, self-re-evaluation, counter-conditioning, helping relationships, reinforcement management stimulus control and self-liberation) that are actually cognitive and behavioral activities [34] that facilitate moving through the stages (table 2).

**Table 2.** Stage of change, techniques and messages.

| Change |  |  |  |
| --- | --- | --- | --- |
| Precontemplation | Raise awareness | Un tratamiento farmacológico adecuado le podría ayudar a prevenir la aparición de nuevos eventos cardiovasculares | An adequate pharmacological treatment could help prevent the appearance of new cardiovascular events |
| Contemplation | Give information and promote personal reevaluation | Recuerde que el evento cardiovascular requiere control y cuidados de por vida | Remember that a cardiovascular event requires lifelong control and care. |
| Preparation | Promote self-care | La EPS es responsable de suministrarle los medicamentos. Exija su derecho | The EPS<sup>1</sup> is responsible for supplying the medications. Claim your right. |
| Action | Offer behavioral guidelines | Si presenta dificultades para recordar la toma de medicamentos pídale a alguien que le ayude a recordarle | If you have trouble remembering to take your medication, ask someone to help you remember. |
| Maintenance | Offer behavioral guidelines | Procure no quedarse sin cobertura de salud, los profesionales en salud pueden darse información de diferentes formas de vinculación | Make sure you do not lose your health coverage. Health professionals can give you information about different types of enrollment. |

### Program design

The intervention was designed for SMS to be delivered over 12 months, based on the stages of the TTM. In the first month, we expect many patients to be in the pre-contemplation phase so messages will be sent daily and aim to raise awareness of future cardiovascular risk. During the second month, the SMS will be sent every other day (four messages per week) and then once per week during the remaining ten months. Thus between 80 and 90 SMS were needed to complete the one-year intervention program. The *CommCare* platform was selected as the mobile platform for collecting data and managing the frequency and timing.

The SMS will be sent through *Telerivet*, an instant SMS platform for organizations as a unidirectional messaging program. The program will not offer interaction with the patients and the messages will not be tailored. If at any time the patients want to withdrawal from the trial, they will have the option to reply with the word STOP. The Abroms’ model recommends clearly identifying the source of the SMS and associating it with the health program [20]. To follow this recommendation and to make it easier for the participants to recognize the message as being part of the study intervention, we designed a step in the randomized trial protocol for saving the contact in the participant’s cell phone, with the name *Proyecto Txt4/FCV (Txt4 Project/ FCV)*.

### SMS Construction

A total of 415 messages were generated by the interdisciplinary group. They classified 212 messages as viable, 116 messages as editable messages and 87 messages to be discarded. The result of this stage was a text message library of 343 SMS to be evaluated by the judges (see Figure 2). The validation by experts resulted in a total of 133 messages that met the criteria of agreement (at least an average of 3.7 for the three aspects and a Kendall’s W of .85 or higher among the judges).

**Figure 2.**
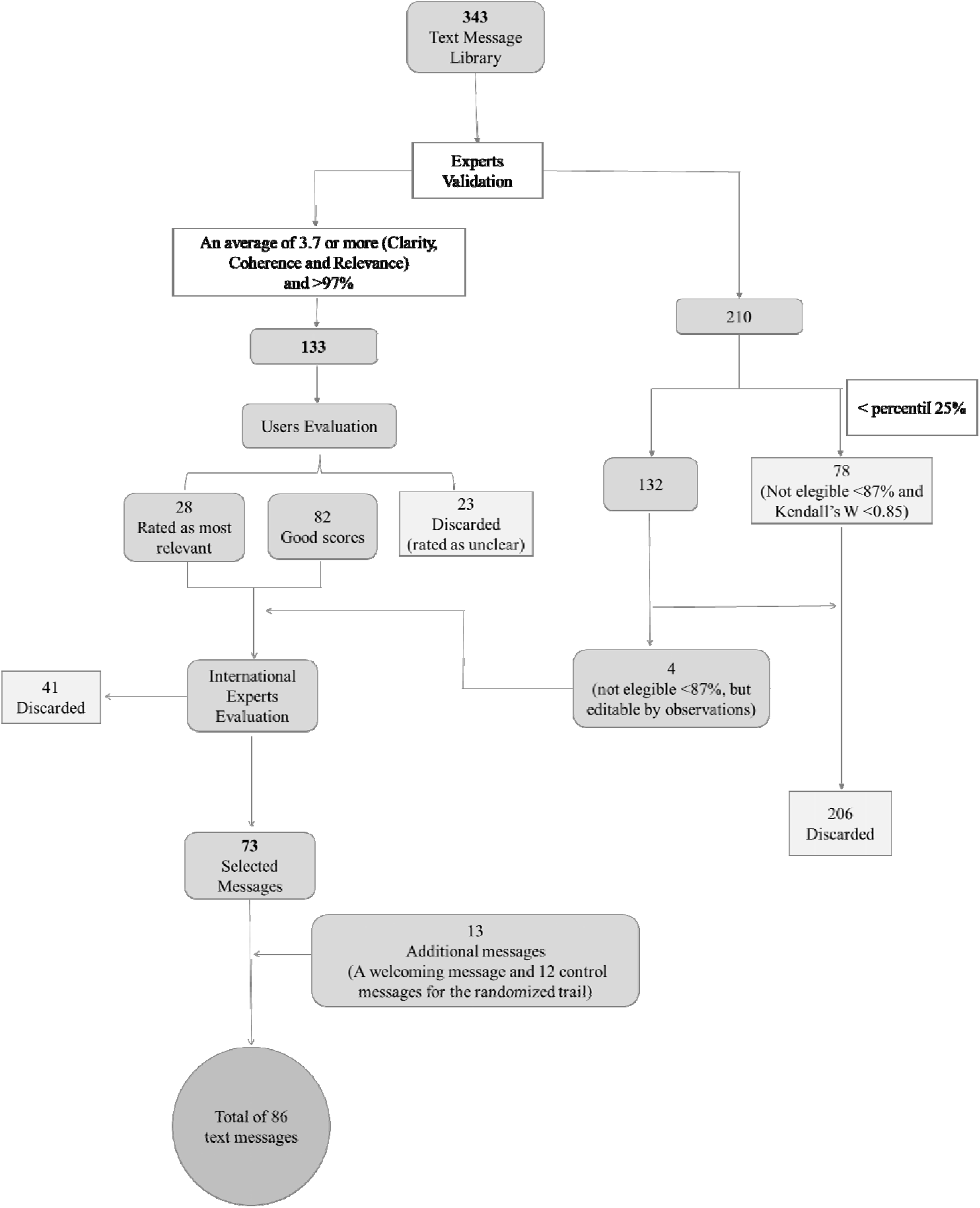
Process to Construct Text Messages.

### Pretesting

Of the 133 messages read by the 9 cardiovascular patients, 110 (83%) were rated as clear, and 28 of those were scored as the most relevant to the patients. Participants also commented on the wording and punctuation of the messages, indicating that they were short and brief, with no technical terms. These 110 messages were presented to the international experts for final review and recommendations. They also evaluated other factors, such as the sequence of the SMS for future trials and the avoidance of repetitive information. A key recommendation was to prioritize messages that referred to medication adherence. Thus, 73 SMS were finally selected for use in the trial (see Table 3). This step also included testing the program configuration. The SMS were successfully sent to and received by the research team’s cell phones.

**Table 3.** Number of messages and distribution.

| Category | Stage of Change |  |  |  |  | Total per category |
| --- | --- | --- | --- | --- | --- | --- |
|  | Pre-contemplation | Preparation | Contemplation | Action | Maintenance |  |
| Disease perception | 2 | 2 | 3 | 2 | 3 | 12 (16%) |
| Health system perception | 2 | 3 | 2 | 5 | 3 | 15 (21%) |
| Supportive networks | 2 | 2 | 2 | 5 | 3 | 14 (19%) |
| Medication intake | 7 | 5 | 7 | 7 | 6 | 32 (44%) |
| General total: | 13<br>(18%) | 12<br>(16%) | 14<br>(19%) | 19<br>(26%) | 15<br>(21%) | 73<br>(100%) |

### Final Revision

The research team made a final evaluation. During this revision, additional messages were created to prepare the program for the randomized trial, especially for the control group. One welcome message was developed and 12 messages for the control group. These were related to recognition and gratitude for participation in the study. Thus, 13 messages were added, but were not related to the intervention for behavior change. Figure 2 summarizes the results of the process.

## Discussion

The objective of this paper is to present the development of an intervention program aimed to increase medication adherence in cardiovascular patients, using technology and a behavior change model. The process was guided by a mixed methods approach that included focus groups with patients, an interdisciplinary team to create the text messages and expert validation. Beginning with the literature search, each step was informed by the findings of the previous one.

Strengths of the approach described include the focus on sustainable behavior change. Hence, the text messages were not reminders for medication intake or appointments, but an intervention to increase awareness and commitment to medication taking. The use of theory (TTM) to inform the content and delivery schedule of the text messages is a strength of this program design, as interventions based on theory have been shown to be more effective than those not using theory (ref), The use of the TTM resulted in a conceptualization of change as a process rather than a one-time event and resulted in a 12-month intervention program with SMS reflecting movement through the stages of change. While the participants are in the first’s stages, motivational and informative interventions are relevant. Therefore, in the program design, the messages sent to the cell phones during the beginning of the intervention, focused on these areas. This action is common for text messaging programs during important behavior change periods [20].As the participants move to subsequent stages, the messages are related to decision making and assuming responsibility. In the action stage, even though the frequency of SMS decreased, the stage has a greater number of messages in comparison with the others, since it aims to develop the adherence behavior. This variation in the frequency, especially decreasing the number of SMS, has been shown to be more effective than a fixed frequency [35].

Another strength of the development strategy was that the SMS were written by an interdisciplinary team of medical specialists and psychologists. This ensured the intervention reflected both clinical knowledge about cardiovascular diseases and the effect of medication and psychological knowledge about cognitive and motivational aspects of behavior change. For example, the physicians developed the messages related to knowledge of the disease, treatment, and the importance of taking medication, while the psychologists offered guidelines about the appropriate way to communicate the information through an SMS, in accordance with the TTM.

The focus groups with cardiovascular patients at the beginning and at the end of the process were important to identify the target population’s perceptions of important barriers to medication adherence and user test the SMS for clarity, acceptability and probable impact in the target population [36]. Expert validation provided additional quality assurance.

### Limitations

Limitations to this research must be considered. Due to the specific study site, sampling, and sample size, generalizability of the results to other settings in Colombia or outside Colombia may be limited. The qualitative approach to data collection provided insight into subjective perceptions about messaging text as a strategy to increase adherence medication in cardiovascular diseases patients, but assessment of actual receptivity and uptake messages program and impact on healthy behaviors requires further evaluation. Finally, because of the changing nature of mobile technology and user expectations in m-health devices, the specific recommendations reported here about a text messaging program for secondary prevention in cardiovascular diseases may have time-limited relevance.

Nonetheless, if proven effective the message content could easily be delivered via a range of digital media such as an app or social media messaging.

## Conclusion

We demonstrated the feasibility of developing unidirectional messaging intervention targeting adherence to medication for secondary prevention of heart disease. Considering the long-term condition of the cardiovascular disease, a SMS intervention can be also contemplated as an accompanying experience using technology. It can be useful as a tool to offer support for the patient, with the conviction it comes from expert, as an incentive from the health system.

## Data Availability

The present work will not generate individual data. We will publish the results in an original article, and we will share full data with anyone who requests it through an email. Interested people will be able to communicate by e-mail with our correspondence author, Anderson Bermon, who will get the approval from coauthors to share the document. In the e-mail, the interested people should indicate their names, affiliation and reasons for wanting our full results.

## Notes

### Competing Interest Statement

The authors have declared no competing interest.

### Funding Statement

This work was supported by COLCIENCIAS code 656672553352 grant 899-2015,
Fundación Cardiovascular de Colombia, Floridablanca,
London School of Hygiene and Tropical Medicine, UK Medical Research Council Funded Reference MR/N021304/1
and Pontifical Bolivarian University, Bucaramanga sectional

### Author Declarations

All relevant ethical guidelines have been followed and any necessary IRB and/or ethics committee approvals have been obtained.

Any clinical trials involved have been registered with an ICMJE-approved registry such as ClinicalTrials.gov and the trial ID is included in the manuscript.

